# Incidence, Clinical Features and Outcomes of Acute Kidney Injury in Adults and Children Admitted with Dengue Infection in Jamaica

**DOI:** 10.64898/2026.03.26.26349368

**Authors:** Tone Wilson, Jerome Walker, Rebecca Thomas-Chen, Lori-Ann Fisher

## Abstract

**Background:** The global burden of dengue infection has rising, yet limited data exists on its impact in the Caribbean. We describe the incidence and associates of acute kidney injury in adults and children with dengue at a teaching hospital in Jamaica.

**Methods:** A single-centre retrospective cohort study of admissions with laboratory confirmed dengue infection at University Hospital of the West Indies, Mona Jamaica between January 2023 to November 2024. AKI was defined using Kidney Disease Improving Global Outcomes definitions. Patients were included if aged >1year and had at least 2 creatinine values. Clinical, demographic and laboratory data were abstracted by chart review. Summary statistics were used to describe continuous and categorical data, and logistic regression to determine AKI associations. Stratified analysis was performed by age-group (adults-aged ≥ 16, and paediatric-aged <16 years).

**Results:** Analyses included 167 persons, 62% (103) were male, mean age was 26.1±19.5 years. AKI occurred in 25.8%, 65.1% were KDIGO stage 1. AKI incidence was 30.2% and 18.0% among adults and children respectively. There were 3 in-hospital deaths. People with AKI were older 32±21.4 vs 24 ±18.4 (p=0.021), and had longer duration of stay [6 vs 4 days (p <0.001)]. Male sex [OR 2.09 (95% CI:0.96-4.59), p=0.064], age per year [OR 1.02 (95% CI:1.01-1.04), p=0.015] symptom duration [OR1.11 (CI 0.99-1.24), p = 0.058], admission bilirubin [OR 1.02 (CI: 1.00-1.04), p = 0.022], NLR [OR 1.09 (CI 1.00-1.18), p = 0.037] were associated with AKI. In adults admission potassium was inversely associated with AKI [OR 0.46 (95% CI 0.21-1.01), p 0.056], while in children admission potassium [OR 3.00 (95% CI 0.88-10.6), p 0.088] was associated with AKI.

**Conclusion:** AKI in dengue hospitalizations is higher than most reports at 25.8%. Targeted public health policy on vector control and early symptom recognition may be needed to improve outcomes.

## Introduction

The year 2024 has been documented as the worst year for Dengue, with an estimated 14 million cases resulting in over 9000 deaths, the highest prevalence and mortality arising from the Americas region(1, 2). Dengue virus has now been classified as a stage 3 WHO Emergency requiring special international attention and support and hence continual global surveillance(2).

Dengue virus is an RNA arbovirus with four (4) main serotypes DENV 1 through 4, transmissible to humans through the bite of an infected female Aedes aegypti mosquito(3). A fifth serotype (DENV 5) was described in 2013 in Malaysia(4). It is endemic to tropical and sub-tropical climates such as the Americas which contains the Caribbean region(2). Dengue is marked by low level ongoing infections underwritten by seasonal bursts often of epidemic proportion termed “peaks/outbreaks”(1, 5). These occur during the “dengue season” which spans the months of March to November annually, with increased hospitalization and case fatalities(1, 2, 6). Increasing disease burden and rising mortality rates has been reported over the last two decades due to increasing case numbers largely effected by urbanization, climate change and resource dependent factors(1-3, 6).

The Americas has a high burden of dengue infection and continues to experience annual dengue outbreaks(1). These occur despite local efforts inclusive of public education and vector control programmes bolstered by social and environmental factors (high population density, poor housing, delayed vector control delivery and delays in and limited access to healthcare and resources particularly in rural areas)(2, 5, 6). Jamaica, a middle income country in the Caribbean(7), is not exempt from these issues, due to both environmental stressors (e.g. hurricanes, increasing temperatures) and rapid urbanization(1, 5, 8). In late November 2023, an estimated 3147 confirmed or suspected dengue cases were reported from the Ministry of Health and Wellness in Jamaica(8).

The dengue illness has a wide spectrum of presentations from asymptomatic, self-limiting acute viral illness to severe illness which spans, single and multi-organ injury and failure (9-11)including acute kidney injury. Renal manifestations of Dengue include rhabdomyolysis(12, 13), proteinuria(14, 15), haematuria, acute tubular necrosis and glomerulopathies(10, 14, 16, 17).

Acute Kidney Injury (AKI) is a common global health problem, with a disproportionate burden in low and middle income countries, in which infections are amongst the predominant aetiologies(18-20). It is independently associated with increased health care costs, mortality and long term risk of chronic kidney disease(20, 21). There have been limited reports on dengue-associated AKI across global literature. A meta-analysis of 37 studies with 21 764 participants reported a pooled prevalence of AKI in dengue of 8% (95% confidence interval 6 to 11), with high heterogeneity across studies due to variability in AKI definitions and study populations. Notably most of the studies were from East Asia and West Africa(22).

Rates of AKI maybe higher in severe dengue, complicating up to a third of such admissions (23). From a meta-analysis of 9 studies, severe dengue, male sex, diabetes and chronic kidney disease were associated with AKI(24, 25). There is a paucity of data on AKI prevalence in children. Rates of AKI amongst paediatric hospitalizations range from 6 to 21%(16, 17), the higher prevalence in those admitted to the intensive care unit with severe dengue infection(16).

Limited data exists on AKI epidemiology in dengue in the Caribbean. From an analysis of community acquired AKI from Jamaica, 4% of emergency room visits had AKI, with viral infections amongst the common causes(26). Understanding AKI risk factors and outcomes may aide in generating public health policy to reduce impact and risk of AKI in the region. We aimed to determine the incidence, associated factors and outcomes of adults and children admitted to a single tertiary hospital in Jamaica.

## Materials and Methods

### Study Design and population

This study is a single-centre retrospective cohort study from January 2023 - November 2024 of Dengue cases at a teaching hospital in Jamaica. Informed consent was waived. Ethical approval attained by Mona Research Ethics Committee at the University of the West Indies Mona Campus with assigned number CREC-MN.0115,2024/2025. The study was conducted between May to December 2025.

Patients were included if they were admitted for at least eight (8) hours within the Emergency Department or any of the medical or surgical floors; are at least age one (1) year and older; had at least 2 accessible creatinine measurements within the prior 1-year period and up to and during hospitalization period, at least one (1) of which was done during the current hospitalization; and had an acute viral illness or dengue-like illness prompting presentation, or onset occurring during hospitalization period with corresponding confirmatory Dengue testing representing an acute or recent infection i.e. positive non-structural(NS1) antigen or immune assay (IgM antibody). Patients were excluded if they were below the age of one (1) year old, had pre-existing End Stage Renal Disease (ESRD) or previously established on renal replacement therapy; had less than two (2) available creatinine measurements within past 1-year or period of hospitalization and were suspected cases of Acute Viral Illness/ Dengue without confirmatory testing i.e. NS1 Antigen or immune assays.

### Procedures

The University Hospital of the West Indies, Jamaica is an academic tertiary referral centre with 547 beds, and has both adult (aged 16 years and older) and paediatric inpatient services(under the age of sixteen years). The hospital was in a transitory period concerning documentation of medical information - from written physical patient notes to electronic notes. The majority of Emergency Department documentation was done electronically via Hospital Information Management System (HIMS). Adult admission documentation was done electronically via the HIMS. Paediatric admission documentation was however still written in paper dockets. All dockets were to be coded at death or discharge as per International Classification of Diseases, 10th Revision, Procedure Coding System (ICD-10-PCS) standardized coding and filed in Medical Records Department Library.

A log of all admissions throughout January 2023 to November 2024 of all ages was retrieved from the UHWI Medical Records Department using ICD-10-PCS codes for Dengue and Acute Viral Illness (as transcribed at death/ discharge). The ICD-10-PCS codes included were: (i) A97.0 Dengue without warning signs; (ii) A97.1 Dengue with warning signs; (27) A97.2 Severe Dengue (Dengue Haemorrhagic); (iv) A97.9 Dengue, unspecified and (v) B34.9 Viral infection, unspecified.

Patient files were reviewed for confirmatory dengue NS1 Antigen and/or immune assays (IgM±IgG) done at / after the point of symptom onset and ordered and reviewed by an external referring physician or done upon/ during hospitalization. Paediatric patients had review of records for confirmatory dengue testing. Adult patients had review of electronic Emergency Department and electronic Internal Medicine admission notes via HIMS for confirmatory dengue testing.

All patients of ages one year and older with confirmatory Dengue testing had laboratory information reviewed from medical records.

Charts were reviewed for age, sex, race, comorbid chronic illness; presenting blood pressure, pulse, respiratory rate and room air oxygen saturations, hydration status, symptoms at presentation, duration of symptoms, medication exposures, need for other interventions (oxygen, blood products, intensive care support); baseline laboratory investigations at admission and discharge. For paediatric charts we obtained arthrometric measurements (height in centimetres and weight in kilograms). If height was not available in the medical record we calculated the height for the 50^th^ centile for age.

### Definition of Variables

1. AKI is defined (based on Kidney Disease improving global outcomes definition (KDIGO) (20)as any of the following:
  a. An increase in serum creatinine (SCr) by x 0.3 mg/dl (X26.5 umol/l) within 48 hours;
  b. or an increase in SCr to X1.5 times baseline, which is known or presumed to have occurred within the prior 7 days;
  c. or Urine volume 0.5 ml/kg/h for 6 hours

According to these criteria, patients were assigned to KDIGO Stage 0 (no AKI), KDIGO Stage 1, 2, 3 based on peak creatinine in the admission compared to the baseline creatinine. If baseline creatinine is unavailable, the peak creatinine was compared to the admission/or lowest creatinine.

Dengue infection –

A. confirmed: positive NS1 Antigen and/or positive IgM serology done at or within 10 days of symptom onset of a dengue -like illness
B. suspected: dengue -like illness without positive NS1 Antigen and/or positive IgM serology done at or within 10 days of symptom onset

2) Dengue-like Illness - acute onset febrile illness accompanied by one of the following: nausea, vomiting, rash, petechiae, aches and pains, leukopenia

3) Acute Viral Illness - acute onset febrile or non-febrile illness associated with malaise, aches, rash, nausea and vomiting

### Measurements

All laboratory measurements were obtained from the UWI Mona laboratories.

Samples for creatinine were analysed using a Cobas c111 Analyzer using a standardized Jaffe method. Estimated Glomerular Filtration Rate (eGFR) was calculated using the 2021 Chronic kidney disease-Epidemiology (CKD-EPI) Collaboration equation(28) for persons aged>=16 years, and Schwartz-Lyon equation for those <16 years(29).

A Cell-Dyn Ruby-Haematology Analyzer was used to analyse blood counts and automated differentials. Neutrophil lymphocyte ratio was determined by dividing the absolute neutrophil count by the lymphocyte count on admission.

Dengue Immunoglobulin M (IgM), IgG Antibody and Non Structural 1 antigen were performed using the Abbott Bioline™ Dengue Duo kit. This is an immunochromatographic assay used to detect NS1 antigen and IgG/M from the serum or plasma.

### Power calculation

Assuming an AKI incidence of 8% in the population (range from 7-16%), our sample size of 167 had a power of 85.2%, with margin of error of 5% to estimate this incidence.

### Statistical Analysis

Data were collected using spreadsheet and exported to the Research Electronic Data capture (REDCap) which is a secure web platform for building and managing online databases and surveys, hosted at the Mona Information Technology Services at the University of the West Indies, Mona campus(30). Data collected were de-identified and stored in a password protected database and exported to STATA software (version SE 17.1; StataCorp LP, College Station, Tex.). Continuous variables were described using means and standard deviations when normally distributed, and medians and interquartile ranges when skewed. Categorical variables were described using frequencies or percentages. Differences in baseline characteristics of AKI were determined by the paired t-test or Mann-Whitney Test whilst differences in proportions were determined via the Pearson’s chi-squared tests or Fisher’s exact test as appropriate. Multivariable logistic regression models adjusting for age and sex were performed to determine AKI associations for the entire cohort. We performed a stratified logistic regression by adult and paediatric groups: adult was defined as age ≥16 years, and paediatric <16 years.

### Outcomes

Cases were examined for primary endpoint of AKI. Secondary end points were death from any cause in hospital, initiation of dialysis and length of hospital stay

## Results

### Cohort Selection

A total of 445 medical records were reviewed for suspicion of dengue and acute viral illness with ICD-10-PCS codes for Dengue and Viral Infection unspecified. Of these, 219 cases had the clinical diagnosis of Dengue at discharge/ death. One hundred and sixty seven (167) patients met eligibility criteria with confirmatory testing based on positive Dengue NS1 Ag and/or immune assay IgM and creatinine results. (see Figure 1 for cohort selection).

**Figure 1:**
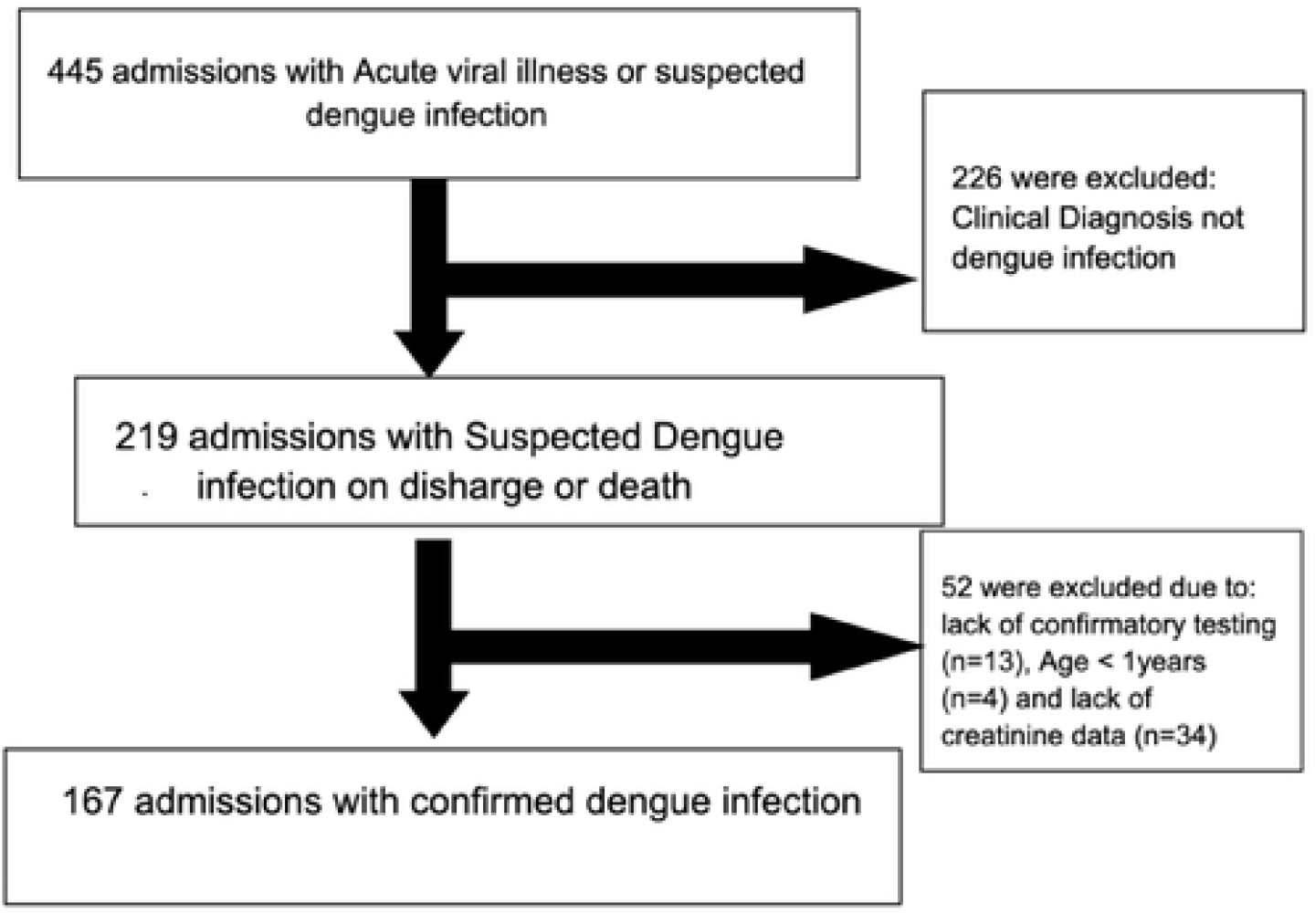
Study cohort diagram. Of 445 admissions with acute viral illness or dengue infection, 219 had clinical diagnosis of dengue infection on discharge or death, 52 were excluded due to lack of confirmatory testing (n=13), age<1 years (n=4), lack of creatinine data (n=34)

### Demographic and Clinical Characteristics of Cohort

Of the 167 participants included, there were 103 (61.7%) males. Age ranged from 1 to 86 years, mean age ± SD was 26.1±19.5 years. Thirty seven percent were in the paediatric age group (aged<16 years). Table 1 highlights baseline differences in the study cohort by sex. For sex differences in adults see supplemental table 1. Diabetes and Chronic Obstructive Pulmonary disease (COPD)/Asthma were the commonest comorbidities in 9.6 and 10.8% of the study population. Median IQR duration of symptoms was 4 (2,5) days. AKI occurred in 43 (25.8%), with no sex differences in AKI incidence (30.8% males versus 18.1% females, p=0.108). There no sex differences in baseline comorbidities, duration of symptoms, admission systolic and diastolic blood pressure, proportion with fever on admission. However females had higher median heart rates on admission (108.5 versus 98 bpm, p=0.036). As expected females had lower mean haemoglobin (12.8 versus 14.1 g/dL, p<0.001) and median creatinine than males (66 versus 87 micromoles/L, p<0.001). However mean eGFR was similar between males and females. There is no difference in admission estimated glomerular filtration rate between males and females.

**Table 1:** Demographic, clinical characteristics of the study population stratified by sex.

| Demographic & Clinical Parameters | All (167) | Male (n=103) | Female (n=64) | p value | Missing (n) |
| --- | --- | --- | --- | --- | --- |
| Mean ±SD Age, (years) | 26.1 ± 19.5 | 24.8 ± 17.5 | 28.0 ± 22.2 | 0.306 | |
| Paediatric, n (%) | 61 (36.5) | 37 (35.9) | 24 (37.5) | 0.837 |  |
| Hypertension, n (%) | 16 (9.6) | 7 (6.8) | 9 (14.1) | 0.121 |  |
| Diabetes, n (%) | 12 (7.9) | 5 (4.9) | 7 (10.9) | 0.139 |  |
| COPD/Asthma, n (%) | 18 (10.8) | 9 (8.8) | 9 (14.1) | 0.281 |  |
| Sickle cell Anaemia, n (%) | 11 (6.6) | 4 (6.3) | 7 (6.8) | 0.890 |  |
| Median (31) Baseline Serum Creatinine (micromoles/L) | 62 (45,76) | 69 (54,80) | 53.5 (41,68) | 0.002 |  |
| Median (31) Duration of symptoms, (days) | 4 (2,5) | 4 (2,5) | 3.0 (2,4.5) | 0.115 |  |
| Mean ±SD Admission Systolic Blood Pressure, (mmHg) | 114.4 ± 17.0 | 113.9 ± 16.9 | 115.1 ± 17.2 | 0.664 | |
| Mean ±SD Admission Diastolic Blood Pressure, (mmHg) | 68.6 ± 11.1 | 67.5 ± 10.9 | 70.2 ± 11.4 | 0.141 | |
| Median (31) Admission Heart rate (beats per minute) | 99 (83,116) | 96 (80,115) | 108.5 (91.5, 120) | 0.036 |  |
| Fever on admission, (Temperature ≥ 38 degrees Celsius) n (%) | 61 (36.5) | 38 (36.9) | 23 (35.9) | 0.901 | |
| Mean ±SD Admission Haemoglobin, (g/dL) | 13.6 ± 2.3 | 14.1 ± 2.2 | 12.8 ± 1.8 | <0.001 | |
| Mean ±SD Admission Haematocrit, (%) | 42.1 ± 6.8 | 43.4 ± 7.0 | 39.8 ± 5.7 | <0.001 | |
| Median (31) Admission WBC, (x10e9/L) | 4.2 (2.7, 7.3) | 4.2 (2.8, 7.6) | 4.2 (2.5, 7.3) | 0.937 |  |
| Median (31) Admission NLR | 2.3 (1.3, 4.8) | 2.1 (1.1, 4.6) | 2.6 (1.4, 4.8) | 0.203 |  |
| Median (31) Admission Platelet, (x10e9/L) | 122 (64,179) | 145 (64, 214) | 108 (62, 160) | 0.121 |  |
| Mean ±SD Admission Sodium (mmol/L) | 135.1 ± 3.9 | 135.5 ± 4.0 | 135.1 ± 3.8 | 0.986 | |
| Mean ±SD Admission Potassium (mmol/L) | 4.1 ± 0.6 | 4.1 ± 0.7 | 4.1 ± 0.6 | 0.772 | |
| Median (31) Admission AST (U/L) | 78 (45, 135) | 83 (48, 145) | 56 (39,122) | 0.088 | 13 |
| Median (31) Admission Total Bilirubin (umol/L) | 9 (5.5,15) | 10 (7,19) | 7 (5,12) | 0.005 | 31 |
| Mean ±SD Admission eGFR (ml/min/1.73m<sup>2</sup>) | 91.9 ±31.4 | 93.2± 38.1 | 89.5 ± 26.1 | 0.541 | |
| Median (31) Admission Creatinine, (umol/L) | 77.5 (59,100) | 87 (64,112) | 66 (55,80) | <0.001 |  |
| AKI, n (%) | 43 (25.8) | 31 (30.1) | 12 (18.8) | 0.103 |  |
| Median (31) Length of Stay (days) | 5 (3,7) | 5 (3,7) | 4.5 (3,7) | 0.898 |  |
| In- Hospital Death, n (%) | 3 (1.8) | 1 (1.0) | 2 (3.1) | 0.308 |  |

**Key: eGFR: estimated Glomerular filtration rate, AKI= Acute kidney injury, WBC white blood** **cell count, NLR neutrophil lymphocyte ratio, AST aminotransferase**

**Table 2.** Clinical and Demographic Parameters for Dengue Cohort stratified by AKI.

| Demographic & Clinical Parameters | All (167) | AKI (n=43) | No AKI (n=124) | p value |
| --- | --- | --- | --- | --- |
| Mean ±SD Age, (years) | 26.1 ±19.5 | 32.0 ± 21.4 | 24.0 ± 18.4 | 0.021 |
| Hypertension, n (%) | 16 (9.6) | 4 (9.3) | 12 (9.7) | 0.943 |
| Diabetes, n (%) | 12 (7.2) | 1 (2.3) | 11 (8.9) | 0.301 |
| COPD/Asthma, n (%) | 18 (10.8) | 5 (11.6) | 13 (10.5) | 0.835 |
| Sickle cell Anaemia, n (%) | 11 (6.6) | 3 (6.9) | 8 (7.0) | 0.905 |
| Median (31) Baseline Serum Creatinine (micromoles/L) | 62 (45,76) | 69 (49,80) | 62 (45,72.5) | 0.177 |
| Median (31) Duration of symptoms, (days) | 4 (2,5) | 4 (2,6) | 4 (2,5) | 0.210 |
| Mean ±SD Admission Systolic Blood Pressure, (mmHg) | 114.4 ± 17.0 | 117.4 ± 19.7 | 113.2 ± 15.8 | 0.178 |
| Mean ±SD Admission Diastolic Blood Pressure, (mmHg) | 68.6 ± 11.1 | 69.5 ± 12.0 | 68.3 ± 10.8 | 0.528 |
| Median (31) Admission Heart rate (beats per minute) | 99 (83,116) | 110 (93,122) | 97 (81, 115.5) | 0.032 |
| Fever on admission, (T≥38 degrees C) n (%) | 61 (36.5) | 19 (44.2) | 42 (33.9) | 0.226 |
| Mean ±SD Admission Haemoglobin, (g/dL) | 13.6 ± 2.3 | 13.7 ±2.6 | 13.6 ± 2.2 | 0.670 |
| Mean ±SD Admission Haematocrit, (%) | 42.1 ± 6.8 | 42.3 ± 7.4 | 42.0 ± 6.6 | 0.799 |
| Median (31) Admission WBC, (x10e9/L) | 4.2 (2.7, 7.3) | 5.6 (3.6,9.9) | 3.7 (2.4,6.5) | 0.001 |
| Median (31) Admission NLR | 2.3 (1.3, 4.8) | 4.4 (2.2,7.4) | 1.9(1.1, 3.9) | <0.001 |
| Median (31) Admission Platelet, (x10e9/L) | 122 (64,179) | 123(67, 173) | 121 (62, 183) | 0.954 |
| Mean ±SD Admission Sodium (mmol/L) | 135.1 ± 3.9 | 135.3 ± 4.7 | 135.0 ± 3.6 | 0.716 |
| Mean ±SD Admission Potassium (mmol/L) | 4.1 ±0.6 | 4.0 ±0.8 | 4.1 ± 0.6 | 0.157 |
| Median (31) Admission AST (U/L) | 78 (45, 133) | 80 (38,145) | 77 (45,133) | 0.930 |
| Median (31) Admission Total Bilirubin (umol/L) | 9 (5.5,15) | 13 (9,30) | 8 (5,13) | <0.001 |
| Mean ±SD Admission serum albumin (g/L) | 36 (31,44) | 36 (31,44) | 38 (34,41) | 0.580 |
| Mean ±SD Admission eGFR (ml/min/1.73m<sup>2</sup>) | 91.9 ±34.1 | 69.8 ± 31.9 | 99.4 ± 31.5 | <0.001 |
| Median (31) Admission Creatinine, (umol/L) | 77.5 (59,100) | 110.5 (85,135) | 70 (56,87) | <0.001 |
| Median (31) Length of Stay (days) | 5 (3,7) | 6 (4,8) | 4 (3,6) | <0.001 |
| In- Hospital Death, n (%) | 3 (1.8) | 1 (0.81) | 2 (4.7) | 0.102 |

### AKI incidence

AKI occurred in 25.8% (43) of the study population. There were no sex differences in AKI rates in the entire cohort (30.1% males vs 18.8% females, p=0.108). Amongst adults, AKI incidence was 30.2% (36.4% males versus 20.0% females, p=0.075), and in children AKI rates were 18.1% (18.9% males, versus 16.7% females, p=0.823). (See Figure 2a-AKI incidence in adults and children). In terms of AKI severity, most had KDIGO Stage I, 65.1 % (28), while 8 (18.6%) had Stage II AKI, and 7 (16.3%) had Stage III AKI. (Figure 2b).

**Figure 2:**
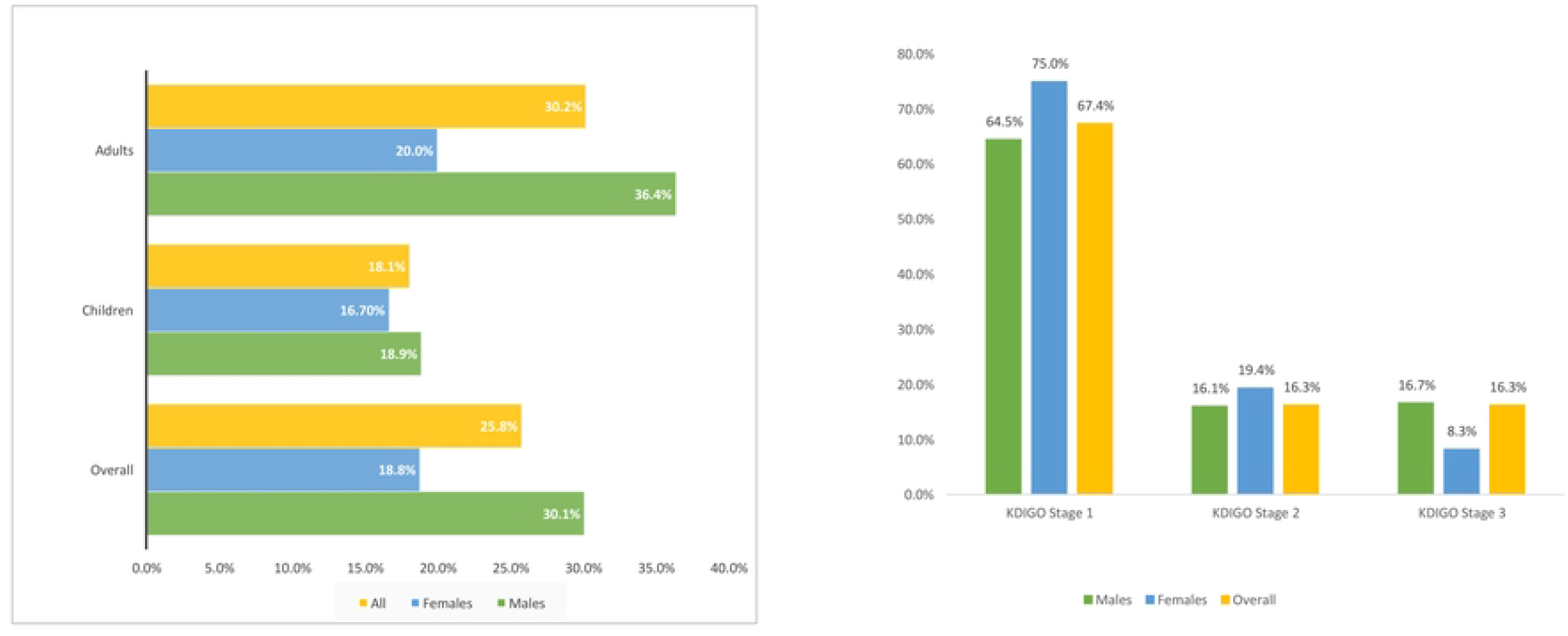
Acute Kidney Injury incidence stratified by (a) Sex and Age group (b) by Kidney Disease Improving Global Outcomes Category.

### AKI Associated Factors

Compared to those with no AKI, those with AKI were older (mean age 32.0 versus 24.0 years, p=0.021), with higher median admission NLR (4.4 versus 1.9, p<0.001), WBC (5.6 versus 3.7, p=0.001) and total bilirubin levels (13 versus 8, p<0.001). There were no differences between frequencies of baseline comorbidities, duration of symptoms, and fever on admission between AKI and non AKI groups. In terms of laboratory values there were no differences between admission haemoglobin, platelet count, sodium, potassium, albumin and aminotransferase levels between AKI and non AKI groups. Median length of stay was higher in AKI than without (6 versus 4 days, p=0.001). For differences by AKI amongst adult and children see supplemental tables 2 and 3.

For results of the logistic regression see supplemental table 1. On multivariable analyses, Male sex [Adjusted Odds Ratio 2.09 (95% CI:0.96-4.59), p=0.064], age per year [OR 1.02 (95% CI:1.01-1.04), p=0.015] symptom duration [OR1.11 (CI 0.99-1.24), p = 0.058], admission bilirubin [OR 1.02 (CI: 1.00-1.04), p = 0.022], NLR [OR 1.09 (CI 1.00-1.18), p = 0.037]. Diabetes mellitus was inversely associated with AKI [OR 0.15 (95% CI 0.02-1.27), p 0.082]. (See Figure 3 For Adjusted Odds ratios)

**Figure 3:**
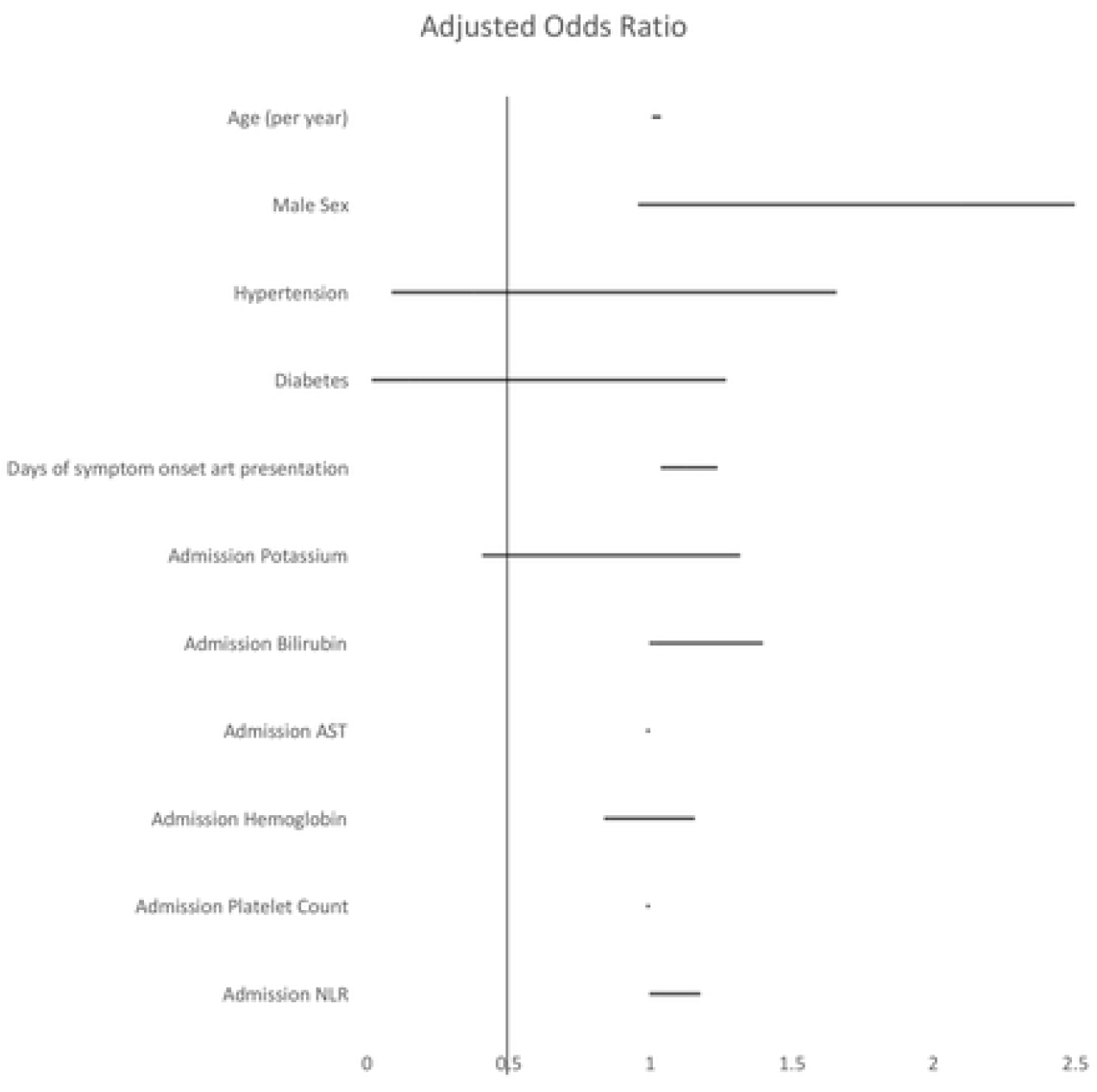
Plots of the with Adjusted Odds Ratios for associated factors and Acute Kidney Injury in the Cohort. Adjusted Odds Ratio: Adjusted for age and sex, AST aspartate aminotransferase, NLR neutrophil Lymphocyte ratio.

#### Associated Factors for AKI in Adults

For differences in characteristics in adults stratified by AKI, see supplemental table 3. A higher proportion of men had AKI (76.7 versus 56.6%, p=0.043). Admission NLR (3.7 versus 2.0, p=0.009), WBC (5.4 versus 3.7 x 10^9^, p=0.05), Total bilirubin (19.5 versus 9 mmoles/L, p<0.001). In multivariable logistic regression analysis for the adult cohort, male sex [aOR 2.68 (95%CI:1.02-7.05), p = 0.045)], admission total bilirubin [aOR 1.02 (95%CI :1.00-1.05, p=0.029), while diabetes [aOR 0.15 (95%CI:0.02-1.86), p = 0.090)] and admission potassium were inversely associated with AKI [aOR 0.48 (95%CI:0.23-1.02), p = 0.056)] (Supplemental table 4)

#### Associated factors for AKI in Children

For differences in characteristics in children stratified by AKI, see supplemental table 2. Of the 61 paediatric admissions, 37 (60.7%) were male, mean ± SD was 9.7 ±4.7 years, 13.5% had asthma and 11.5% had sickle cell anaemia. There was no differences in age, median duration of symptoms, sex, admission vital signs, haemoglobin and platelet count between AKI and non-AKI groups. However, children with AKI had higher admission median WBC (5.1 versus 3.9, p=0.05), and NLR (3.7 versus 2.0, p=0.009) compared to those without. Children with AKI had longer median length of stay than those without (7 versus 5 days, p<0.001). In a logistic regression analysis adjusting for sex and age, admission NLR [aOR 1.40 (95%CI:1.07-1.84), p = 0.014)] and admission potassium [aOR 3.00 (95%CI :0.85-10.6, p=0.088) were associated with AKI. (Supplemental table 6)

## DISCUSSION

In this single-center retrospective cohort study of AKI in hospitalizations with dengue from Jamaica, we found a high rate of AKI (25.8%), with the majority being KDIGO Stage I. Overall male sex, higher bilirubin and NLR on admission and longer symptom duration were associated with increased odds of AKI. In adults while admission potassium and diabetes were inversely associated with AKI; admission potassium and NLR was associated with AKI in pediatric admissions.

AKI in dengue can be multifactorial(2, 6, 10, 32), and there is limited data on mechanisms of renal injury. Volume depletion/ dehydration is a primary cause of AKI, due to insensible losses from fever, vomiting and poor oral intake. Viremia induces release of pro-inflammatory cytokines (inclusive of tumor necrosis factor-alpha, Interleukin-6, 8 and 10), complement activation, and endothelial dysfunction, which results in plasma capillary leak and vasodilation(10, 25, 33). Consequent to this, hemodynamic instability with hypotension and shock may occur resulting in reductions in renal perfusion and tubular injury. Direct viral cytopathic effects on glomerular and tubular cells, and viral invasion into muscle causing rhabdomyolysis may also lead to tubular dysfunction. Glomerular damage due to podocyte injury, and in-situ immune complex deposition may lead to proteinuria and acute glomerulonephritis respectively(10, 24, 33, 34).

Our AKI rates are relatively higher at 25.8%, with high rates of AKI in adults and children. Prior reports of AKI incidence in dengue hospitalizations range from 0.9-69.4%, likely reflecting heterogenicity in definitions of AKI and study populations, some studies limiting analyses to ICU or dengue hemorrhagic fever, in which higher acuity of illness results in higher rates of AKI(9, 14, 16, 17, 22, 25, 35). These differences are more significant amongst pediatric populations in which prior reports of AKI was as low as 6%(17). We used the KDIGO based definition of AKI, which may explain the higher-than-expected AKI incidence amongst pediatric population. Based on a more recent metanalysis, the estimated pooled prevalence rate of dengue associated AKI was 8%. However, there was some variability in AKI prevalence by region, with higher prevalence rates of 13 – 21% for example in Pakistan and Burkina Faso, compared to other territories(14, 22, 23). The variability of AKI prevalence could be explained by differences in virulence of the dengue virus in dengue outbreaks(36), study populations, health-care systems, and access. Our rates of AKI may reflect these factors, in addition to delays in healthcare access, notably longer duration of symptoms was associated with AKI in our analyses(1, 2, 5, 6, 33). Public health policy targeting education symptom recognition, coupled with studies to determine facilitators and barriers of AKI recognition may be needed.

Male sex was associated with AKI in our cohort, concordant with prior observational studies suggesting a high rate of AKI in dengue in males, pooled prevalence of 17% (95% CI 8 to 26) for males compared to 3% (95% CI −2 to 7) for females(22). Murine models purport a modulatory impact on inflammation within rodent kidneys with a more deleterious effect of testosterone on the modulation of mitochondrial protein expression and hence function, as well as a more pro-inflammatory cascade within rodent kidneys with testosterone exposure in comparison to a protective effect of oestrogen in females(37).

Neutrophil-Lymphocyte Ratio (NLR) has been emerging as an indicator of disease severity and prognosis in infections and inflammatory disease (38-40). Few studies have explored the predictive value of NLR in Dengue. In the acute febrile phase of dengue infection higher neutrophil counts (reflecting innate immunity) and hence NLR are expected, however as neutrophil apoptosis increases with resultant lymphocytic predominance, the NLR ratio decreases(41). Prior reports of associations with NLR and dengue outcomes have been discordant. Boer et al, in a cohort study of 193 patients found no association with admission NLR and dengue hemorrhagic fever(42), whilst other observational studies showed positive correlation with severe dengue(43, 44), or moderate diagnostic accuracy and high sensitivity for plasma leak as determined by point of care ultrasound(43). In a metanalysis of 37 studies with 5925 participants, elevated CRP, AST, IL-8 and decreased serum albumin were associated dengue hemorrhagic fever, while vascular cell adhesion protein-1, syndecan-1, AST and CRP were associated with severe dengue infections(24). These findings may highlight the role of early acute inflammation and increased capillary permeability, with hepatic involvement (as measured as increased transaminases) in dengue severity. The finding that NLR (an acute inflammatory marker) and bilirubin (a marker of hepatic dysfunction) were associated with AKI development may also support this. Although we did not access overall disease severity, AKI, is a distinct marker of organ-specific injury(18). That admission NLR was associated AKI, may imply its use for prognostication in dengue infection and may have a role for detecting both systemic and renal inflammation.

NLR is cheap and readily available in low-income settings, and therefore its utility as marker for adverse events should be further explored, validated in other cohorts.

Both hyperkalemia and hyperkalemia have been described in dengue infection(9, 10). Transient transcellular shift due to increase in circulating catecholamines, and/or increased tubular excretion of potassium due to direct viral cytotoxic effects are proposed mechanisms of hypokalemia(45). Contrastingly, hyperkalemia may result from both reduced renal excretion of potassium, and increased cell lysis from hemolysis or rhabdomyolysis(10, 12). The incongruent associations of AKI with potassium in adults and children, may be due to differences in clinical presentations, and degree of tubular dysfunction in adults versus children and could be explored in further studies.

AKI in Dengue infection has high morbidity and mortality rates. Like prior published reports, we showed an association of AKI with increased length of hospitalization(9, 10, 22, 23, 35). Prolonged length of stay adds to the increased health-care burden and both direct and indirect healthcare costs due to costs of hospitalization itself, and loss of productivity. Therefore, efforts to improve recognition and identification of AKI may be needed to improve outcomes in dengue in the Caribbean.

We had a low overall in-hospital mortality, and no persons that required renal replacement therapy. This is likely consequent to the overall younger cohort (mean age was 26 years), less medical comorbidities, and higher rates of mild (KDIGO Stage 1) AKI, all factors which reduce mortality and need for renal replacement therapy in AKI. Interestingly, diabetes was protective for AKI amongst adults, contrary to prior reports of dengue associated AKI. This unexpected result may be due to reverse causality, as patients with diabetes may have had lower thresholds for admission, more aggressive care to reduce AKI risk (hydration, discontinuation of nephrotoxic drugs). This finding could be confirmed in larger prospective studies.

Although this is the first study to estimate AKI incidence in both adults and children in the Anglophone Caribbean, our study has several limitations. Firstly, we did not use urine output criteria to define AKI, which may have underestimated AKI, especially in pediatric populations. Secondly this was a retrospective single center study at a tertiary center which may have limited generalizability, and we did not capture data on disease severity, thereby limiting associations with AKI.

## CONCLUSION

AKI incidence amongst admissions with dengue was high 25.8%, and associated with prolonged hospitalization, further studies should explore AKI risk factors and long-term outcomes of dengue hospitalizations on renal function. Public health policy targeting early symptom recognition are needed to improve outcomes.

## Data availability statement

Data for this manuscript are available on reasonable request.

## Acknowledgements

We would like to thank Professors Michael Boyne on their appraisal of the data analysis and writing of the manuscript. We would also like to thank the medical technologists and administrative staff at the University of the West Indies for their input in this research.

## Authors contributions

TW conceptualized the study, wrote the first draft of this manuscript and collected the data. LAF conceptualized the study, supervised the final analysis, edited and redrafted the manuscript. All authors contributed to editing and redrafting the manuscript.

